# Neurological examination for cervical radiculopathy: a scoping review protocol

**DOI:** 10.1101/2023.05.22.23290194

**Authors:** Marzia Stella Yousif, Giuseppe Occhipinti, Filippo Bianchini, Daniel Feller, Annina Schmid, Firas Mourad

## Abstract

**Introduction:** Cervical radiculopathy (CR) is a common cause of pain in the neck and arm region, with a considerable impact on a person’s physical functioning, mental health, and social participation. The current knowledge of CR is mainly based on empirical concepts and early studies. Although action potential conduction slowing or block of a spinal nerve or its roots (i.e., loss of sensory and/or motor function) is a core sign of CR, among guidelines CR is still commonly defined by pain (i.e., gain of function) radiating into the arm. There is no consensus about the gold-standard for the diagnostic procedures for CR but it has been suggested that clinicians should assess CR by subjective and physical examination including a neurological examination, neural mechanosensitive testing and provocative manoeuvres. Among the several clinical tests routinely used to identify loss of function, the neurological examination historically played a role in the differential diagnosis and in the prognostic profile of radiculopathy. However, there is a paucity of studies investigating the diagnostic accuracy of the neurological examination for CR. Thus, the assessment of CR remains a clinical challenge among primary care clinicians. This often leads to an increased risk of misdiagnosis and inappropriate treatment, potentially contributing to delayed recovery and poor health outcomes. Therefore, according to the population, concept, context (PCC) strategy, our scoping review aims to investigate the evidence in regard to diagnostic accuracy (C) of the neurological examination for CR (P) and raise awareness among clinicians on how to appropriately perform this testing.

**Methods and analysis:** This scoping review will be conducted according to the Preferred Reporting Items for Systematic Reviews and Meta-Analyses (PRISMA) extension for Scoping Reviews checklist and the Joanna Briggs Institute (JBI) Reviewer’s Manual on scoping reviews. The aim of this scoping review is to explore what is already known about the neurological examination and its diagnostic value for CR.

**Ethics and dissemination:** This scoping review will not require ethical approval since it will synthesize information from publicly available studies. Results will be submitted for publication in a peer-reviewed journal, presented at relevant conferences in the field and disseminated through working groups, conferences, webinars, social media.

**Strengths and limitations of this study:**

- Our review will identify knowledge gaps to inform future research about the neurological examination for CR. Diagnostic values will be reported when available.
- To the best of the authors’ knowledge, this is the first scoping review to provide a comprehensive overview on the neurological examination for CR.
- The results will add meaningful information for clinicians to inform assessment of CR. It will also direct future research.
- A robust clinical recommendation might be limited due to the lack of available literature

## Introduction

Peripheral nerve compression and/or irritation within narrow anatomical spaces are known as entrapment neuropathies.^1^ One entrapment neuropathy affecting the upper quadrant is cervical radiculopathy (CR). The etiology of CR remains largely unknown.^1^ However, it is suggested that CR is associated with reduced space for the nerve root in the intervertebral foramen, often triggered by a disc herniation or degeneration causing foraminal stenosis.^1, 2^ Our understanding of CR is still based on early studies, reporting a heterogeneity of pathomechanisms and various clinical presentations. As a consequence, diagnosis and management are challenging for clinicians and epidemiological data are sparse.^1,3-7^ However, CR prevalence was estimated to range from 1.07 to 1.76 per 1,000 and 0.63 to 5.8 per 1,000 for males and females, respectively.^8^ There is variability which is mainly attributable to the differing diagnostic criteria, the geographical population location, and occupational features.^8^

The definition of CR is not universally accepted among guidelines which commonly define CR by pain and symptoms (e.g., paraesthesia) radiating into the arm.^3-7^ However, pain, paraesthesia, heightened nerve mechanosensitivity, hyperalgesia, allodynia and dysesthesia have been described as indicators of neuropathic/radicular pain (namely, gain of function).^9^ Instead, action potential conduction slowing or block of a spinal nerve or its roots is a core sign of CR and it clinically manifests with dermatomal sensory loss, myotomal weakness, and/or deep tendon reflex changes (namely, loss of function).^10, 11^ There is no consensus about the gold-standard for the diagnostic procedures for CR. It has been suggested that clinicians should perform a subjective and physical examination including a neurological examination, neural mechanosensitive testing and provocative manoeuvres.^12^ Notably, the commonly used provocative manoeuvres (e.g., Spurling’s test, axial traction), nerve mechanosenstivity tests (i.e., upper limb neurodynamic tests) and neuropathic pain screening tools (e.g., LANSS, painDETECT, and the stEP tool) have limited clinical use in identifying loss of nerve function^1, 7, 13-17^, but are designed to detect neuropathic/radicular pain.^7^ Indeed, radicular pain from nerve root compression can exist as a discrete disorder without a clinically detectable loss in nerve conduction. To diagnose CR, signs of neurological deficits have to be present.^10^ Diagnostic imaging (e.g., Magnetic Resonance Imaging (MRI)), neurophysiological testing (e.g., electromyography), or surgical verification were also suggested to optimize diagnostic accuracy.^18, 19^ However, the clinical significance of findings on cervical MRI and neurophysiological testing remains contentious due to the high frequency of false positive and false negative results.^1, 18, 20^

Among the several clinical tests routinely used to identify loss of function, the neurological examination for the assessment of peripheral sensory and motor responses has historically played a role in the differential diagnosis and in the prognostic profile of radiculopathy.^18, 21-24^ However, there is a paucity of studies investigating the diagnostic accuracy of the neurological examination for CR.^7, 25, 26^ Thus, the assessment of CR remains a clinical challenge among primary care clinicians. This may lead to an increased risk of misdiagnosis and inappropriate treatment, resulting in delayed recovery and poor health outcomes.^25^ Therefore, this scoping review aims to explore the current evidence of the diagnostic accuracy of the neurological examination for CR, and raise awareness among clinicians on how to appropriately perform this testing.^21, 25^

## Methods and analysis

The proposed scoping review will be performed following the 6-stage methodology suggested by Arksey and O’Malley.^27^ It will be conducted following the extensions to the original framework recommended by the Joanna Briggs Institute methodology (JBI) for scoping reviews and the guidance written by members of the JBI to develop a robust and detailed scoping review protocol.^28^ The 6-stages are as follows: (1) identification of the research question, (2) identification of relevant studies, (3) selection of studies, (4) charting of data, (5) summary of results and (6) consultation with stakeholders, an optional but recommended evaluation component of a scoping review. The PRISMA extension for Scoping Reviews Checklist for reporting will be used to report the final manuscript.^29^ We will follow the framework of Population, Concept and Context (PCC) proposed by The Joanna Briggs Institute to describe the elements of the study inclusion criteria.

### Stage 1: Identifying the research question

We formulated the following research questions:

1. What is known from the existing literature about the evidence regarding the performance and accuracy of the neurological examination in CR?
2. What is reported in the literature about the diagnostic value of the neurological examination in CR?

In particular, the objectives of this study will be to:

1. Provide a comprehensive overview of all studies dealing with diagnostic accuracy of the neurological examination for CR.
2. Inform clinicians on how the neurological examination is performed, according to what is already known in literature.
3. Identify any knowledge gaps regarding the neurological examination and diagnostic accuracy values to inform future research.

### Stage 2: Identifying relevant studies

All named authors have participated in an iterative process to develop the initial search strategy. This includes identifying key terms, inclusion and exclusion criteria and relevant databases.

### Inclusion Criteria

In a scoping review, the three elements of population, concept and context are used to establish inclusion criteria.^28^ The population details the relevant characteristics of participants, the concept is the principal focus of the review, and the context describes the setting under examination.

To achieve transparency and replicability of the full process, the research team will determine the inclusion and exclusion criteria prior to screening. These criteria will be based on the research questions and through discussion of the results from the trial search.

Reasons for exclusion of full-text studies that do not meet the inclusion criteria will be recorded and reported in the final scoping review report. Exclusion reasons will be identified by the authors, based on insights from the trial search, inclusion and exclusion criteria, and observations from the title and abstract screening process.

### Population

This review will consider CR.

### Concept

This review will include studies that report on the performance of the neurological examination and/or its diagnostic accuracy for CR.

### Context

This review will consider studies conducted in any context. Studies that do not meet the above stated PCC criteria or provide insufficient information will be excluded.

### Types of evidence sources

We will include the following primary studies: cross–sectional studies, case–control studies, and randomized controlled trials (RCT) that aim to study the diagnostic accuracy of the neurological examination for CR.^30^ Also, in line with the characteristics of a scoping review, we will include narrative synthesis, systematic reviews, and scoping reviews.^28^ No restrictions regarding time, location, language, or setting will be applied.

### Search strategy

The research group will develop a three-step approach.

1. A preliminary search in PubMed was undertaken to identify articles on the topic and the shared terminology. We analysed all the terms reported to describe the three domains of PCC of interest (Table 1). Variants of the terms identified in table 1 were refined to create a second search strategy with search phrases and Medical Subject Headings (MeSH) terms. The information gained from the initial search was used to develop a more comprehensive search strategy based on the PCC framework for PubMed. Table 1 shows the initial search strategy.
2. A final comprehensive search will be conducted on PubMed, Embase, Scopus, Cinhal, DiTA. The search strategy will be adapted for each database.
3. In addition, grey literature (e.g., Google Scholar) and the reference list of included studies will be searched manually through forward and backward citation tracking strategies (Web of Science) to identify any additional studies that may be relevant to this review.

**Table 1.**
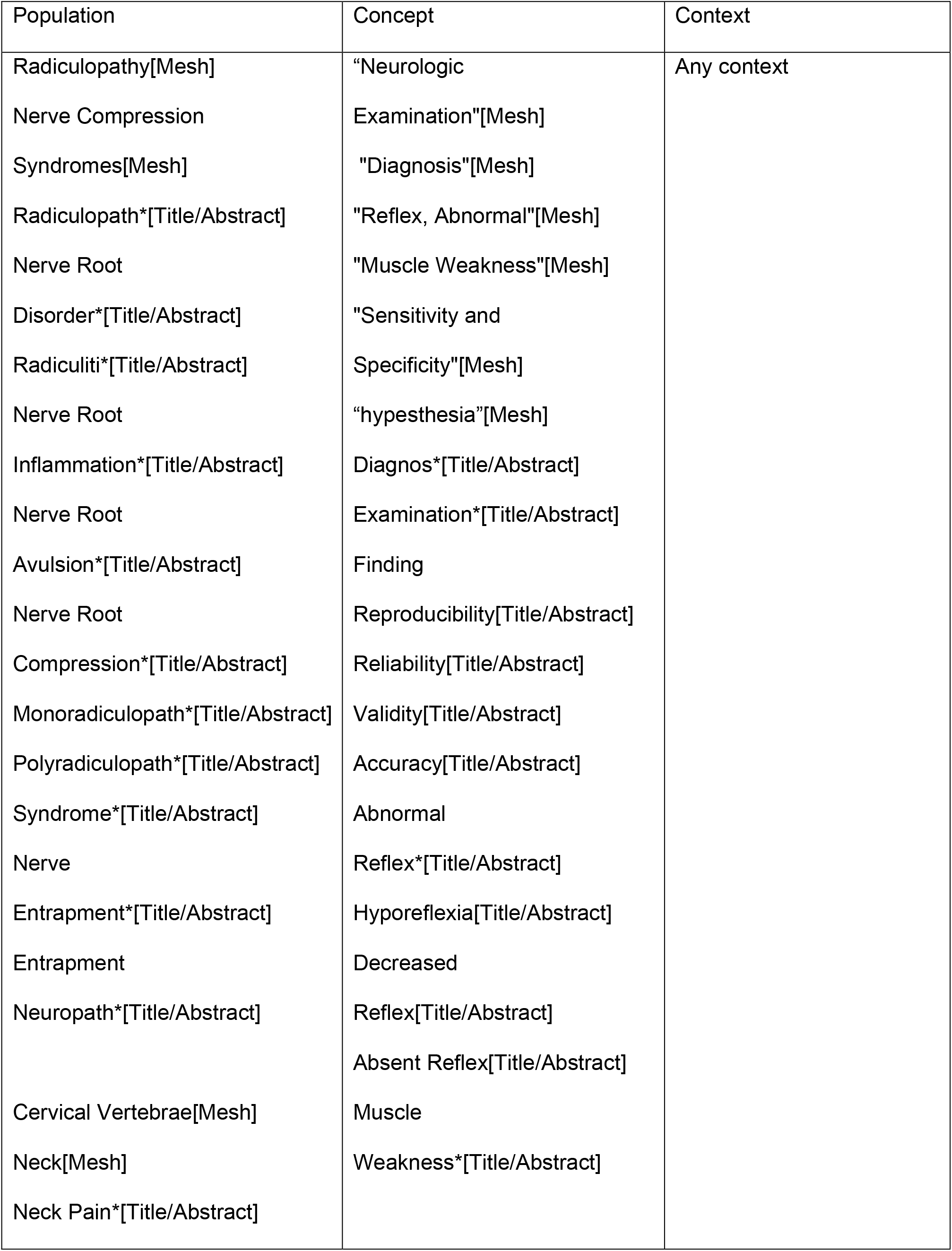
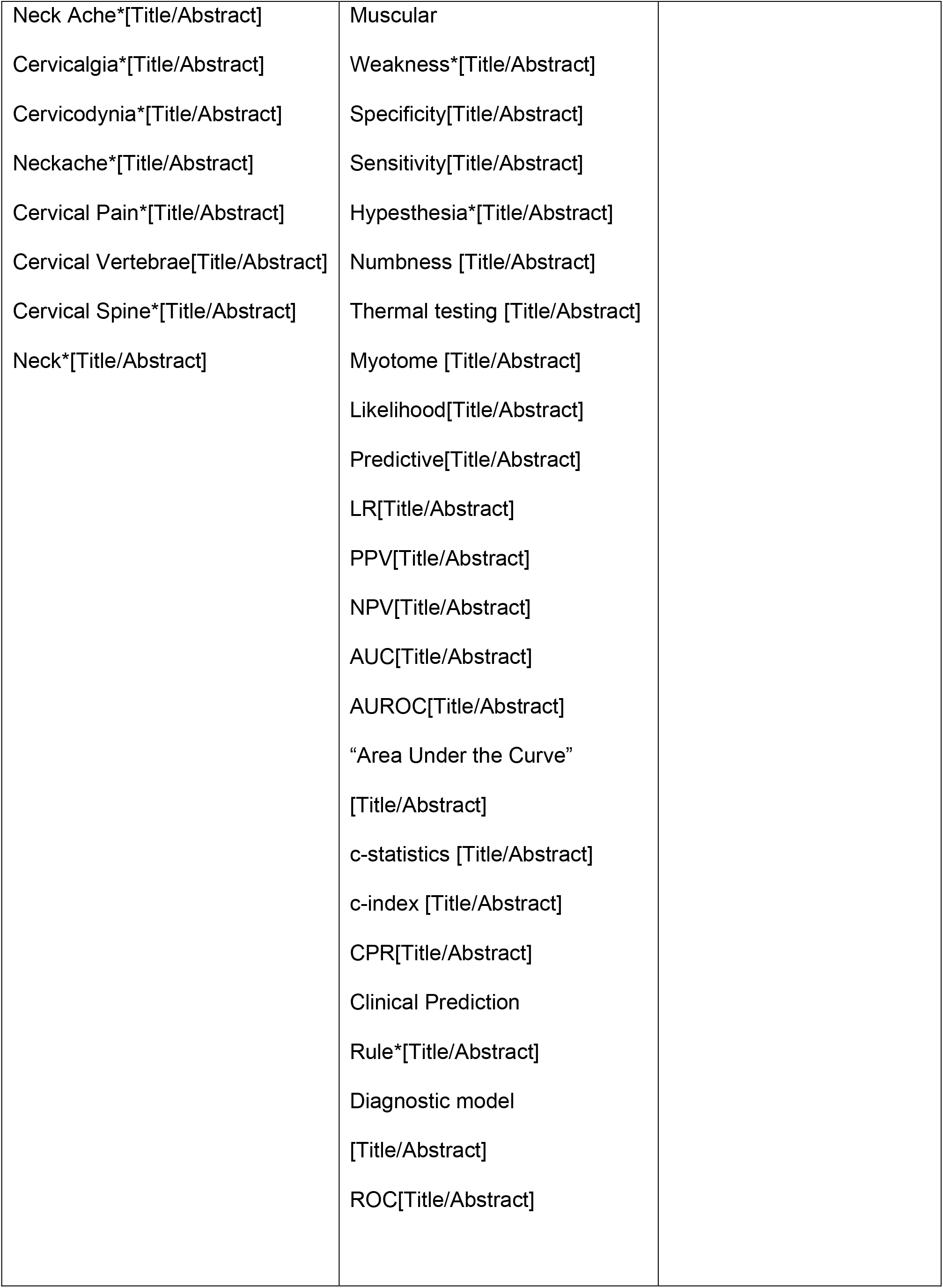
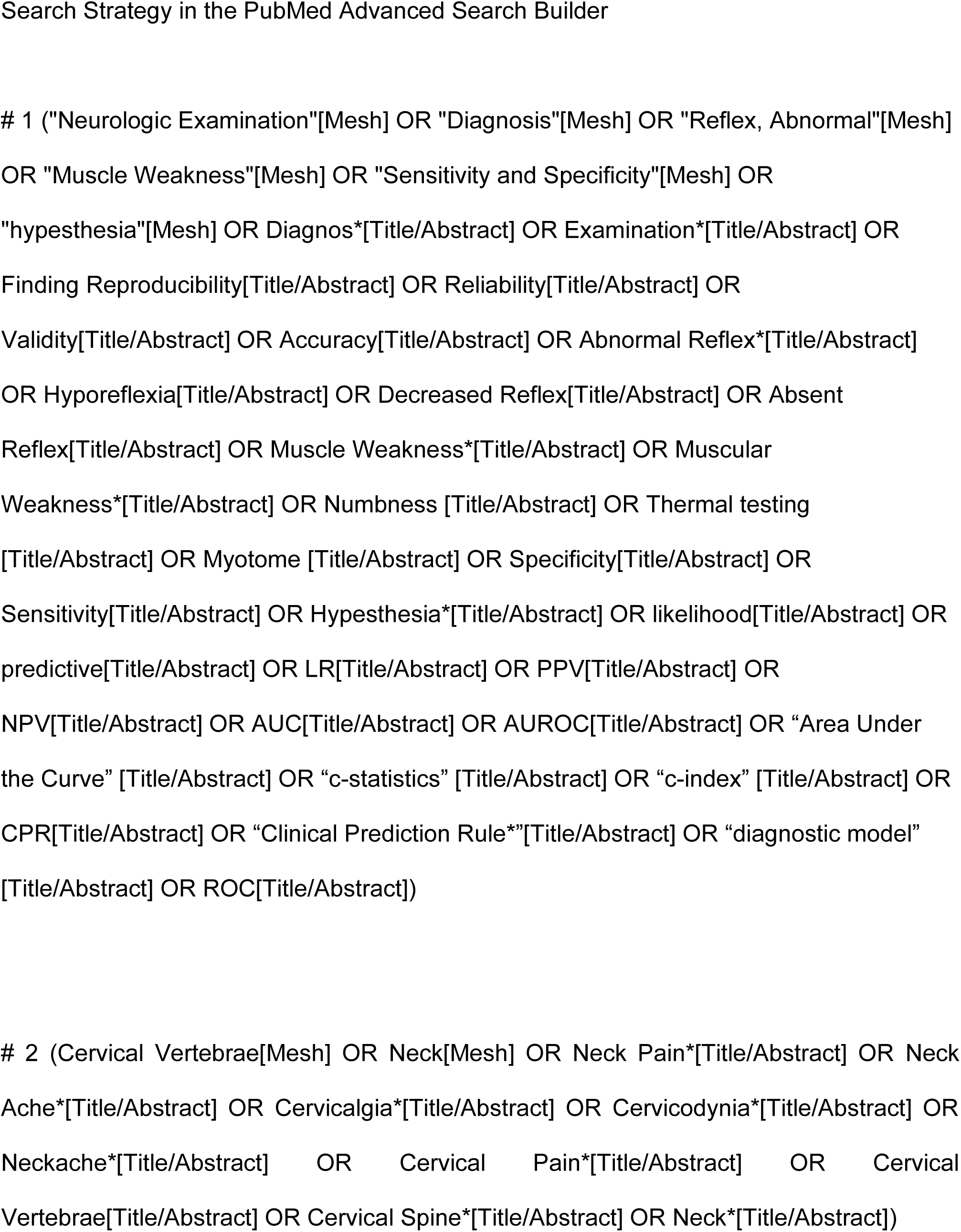
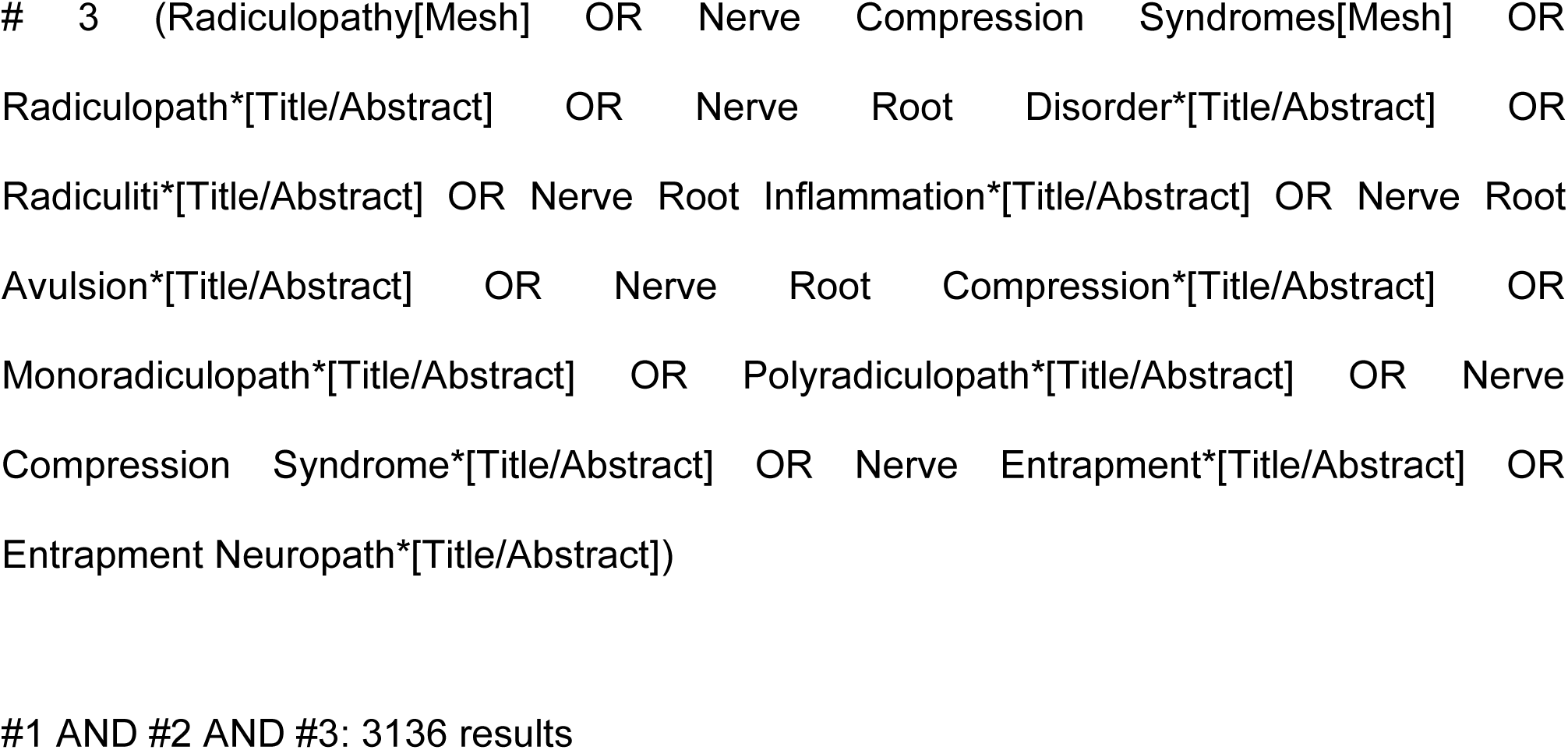
PCC search strategy (PUBMED/MEDLINE)

The PRISMA literature search extension will be used to report the search strategies.^31^

### Stage 3: Study selection

The search results will be collated and exported to EndNote (Clarivate Analytics, PA, USA). Duplicates will be automatically removed before the file containing a set of unique records is made available to reviewers for further processing (i.e., study screening and selection). The review process will consist of two levels of screening using Rayyan QCRI online software: (1) a title and abstract review and (2) a full-text review.^32^ Two investigators will screen the articles independently for both levels to determine if they meet the inclusion/exclusion criteria. In case of any disagreement on inclusion, both reviewers will review the full-text articles again. If agreement cannot be reached, an independent third reviewer will arbitrate. Reasons for the exclusion of any full-text source of evidence will be recorded and reported. The results of the study selection will be reported in full in the final manuscript and summarised in a PRISMA flow diagram (Figure 1).^33^

### Stage 4: Charting the data

The research team will develop a data collection form to extract the characteristics of the included studies. The final sample of articles will be extracted equally and independently among the two reviewers. The extraction tool will be used to collect and chart data. The two reviewers will meet to resolve conflicts, hone their shared understanding of the extraction method and refine the tool when needed. A third researcher will resolve unreconciled disputes. Any modifications to the data extraction strategy will be reported in the results section of the final scoping review.

The initial data-collection form will include the following elements:

- Author(s), year of publication, study location
- Study Population and size
- Aims of the study
- Study design
- Reference test to diagnose CR
- Details of the neurological examination of the peripheral nervous system (i.e., deep tendon reflexes, light touch, muscle strength, pinprick, heat and cold sensitivity), including information on its performance
- Diagnostic accuracy results related to the neurological examination
- Important results and considerations

Missing data will be gathered by contacting the corresponding author with a maximum of two attempts.

### Stage 5: Collating, summarizing, and reporting the results

Results from the scoping review will be summarized descriptively through tables and diagrams. Specifically, we will summarize and synthesize the data extracted from the studies and highlight any knowledge and implementation gaps for the neurological examination in CR. The objectives of each selected study, the concepts or approaches adopted in each article, and the results related to the study research questions will be summarized and explained.

### Stage 6: Consultation with stakeholders

This will be the last step of the review. After analysing and interpreting, preliminary results will be presented to a group of clinical experts and researchers identified by the study team to have expertise in CR or neurological testing (e.g., by the number of publications on the topic). This strategy aims to share and refine preliminary study findings (knowledge transfer and exchange), obtain potentially relevant studies not included in the initial search and develop effective dissemination strategies and directions for future studies.^34^

### Patient and public involvement

This protocol was developed without patient involvement. Patients were not invited to comment on the protocol design and will not be consulted to synthesize outcomes or interpret the results. Patients will not be invited to contribute to the writing or editing of this document for readability or accuracy. The results of this scoping review may inform the development and design of future research study for which patient and public partnership will be sought.

## Data Availability

All data produced in the present work are contained in the manuscript.

## Ethics and dissemination

This study does not require ethical approval as we will not collect personal data. The results will be submitted for publication in a peer-reviewed journal, presented at relevant conferences in the field and disseminated through working groups, webinars and social media. The researchers also intend to form recommendations for areas of future research.

## Contributors

All authors designed the protocol, reviewed the manuscript, approved the final version and participated in the 6 stages.

